# Independent contributions of functional class, comorbidity, and frailty to health status in elderly adults with heart failure

**DOI:** 10.1101/2025.10.21.25337981

**Authors:** Hoang Le Huy Nguyen, Anh Thi Ngoc Pham, Tuan Cong Trinh, The Ha Ngoc Than, Tien Ngoc Hoanh My Nguyen, Tung Huy Pham

**Affiliations:** School of Medicine, University of Medicine and Pharmacy at Ho Chi Minh City, Ho Chi Minh City, Vietnam; Department of Geriatrics and Palliative Care, University Medical Centre Ho Chi Minh City, Ho Chi Minh City, Vietnam; Department of Radiology, Thong Nhat Hospital, Ho Chi Minh City, Vietnam

**Keywords:** Heart failure, Patient-reported outcome measures, Frailty, Comorbidity, Left ventricular ejection fraction

## Abstract

**Aims:** This study identified and quantified independent predictors of health status among older ambulatory adults with chronic heart failure in an Asian population, hypothesizing that systemic burden indicators would predict health status independently of ejection fraction.

**Methods:** This cross-sectional observational study enrolled 150 consecutive Vietnamese patients aged 60 years or older with chronic heart failure attending a cardiology clinic between August 2024 and March 2025. Health status was assessed using the validated Vietnamese Kansas City Cardiomyopathy Questionnaire-12. Multivariable linear regression models examined associations between KCCQ-12 scores and clinical variables including left ventricular ejection fraction, New York Heart Association functional class, Clinical Frailty Scale scores, Charlson Comorbidity Index, and demographics. Secondary analyses evaluated domain-specific scores and phenotype-stratified associations.

**Results:** The median age was 75 years, and median KCCQ-12 summary score was 68. Multivariable regression explained 77% of variance in health status. Advanced NYHA functional class, higher comorbidity burden, and frailty independently predicted lower KCCQ-12 scores, while male sex was associated with higher scores. Left ventricular ejection fraction and N-terminal pro-B-type natriuretic peptide showed no significant association with health status. These patterns remained consistent across all KCCQ-12 domains and heart failure phenotypes. Nutritional status correlated with health status univariately but was subsumed by frailty in multivariable models.

**Conclusions:** Geriatric syndromes, including frailty, multimorbidity, and functional limitation, determine patient-reported health status in older Asian adults with heart failure independent of cardiac-specific indices. These findings support integrating systematic frailty assessment and functional preservation into routine cardiovascular care for aging Asian populations.

## Introduction

Heart failure affects an estimated 64 million individuals globally,^1^ with substantial regional variation in disease burden and outcomes. The absolute number of prevalent cases has increased across all Asian regions from 1990 to 2019, reflecting population aging and improved survival.^2^ This epidemiologic shift has prompted clinical guidelines to emphasize routine monitoring of patient-reported health status using validated instruments,^3^ recognizing that enhancing functional capacity and quality of life are now critical management objectives alongside mortality reduction.^3^ The Kansas City Cardiomyopathy Questionnaire (KCCQ) and its abbreviated 12-item version have demonstrated robust predictive validity for hospitalizations and mortality in diverse settings,^4^ establishing them as preferred disease-specific health status measures in contemporary practice and research.^5^

Patient-reported health status correlates more strongly with clinical outcomes than traditional cardiac metrics, a phenomenon particularly evident in the discordance between left ventricular ejection fraction and symptom burden.^6^ Recent machine learning models demonstrate that KCCQ scores dominate predictions of 90-day hospitalizations and mortality when compared with conventional clinical variables.^7^ Emerging evidence identifies functional class, multimorbidity, frailty, and nutritional status as key determinants of health status in heart failure populations.^8^ Frailty affects at least half of heart failure patients^9^ and independently predicts all-cause mortality,^9^ with nutritional status and frailty demonstrating synergistic prognostic effects.^10^ These systemic indicators often outperform ejection fraction in predicting patient-centered outcomes.^6^

Most predictive frameworks for health status derive from Western cohorts, yet Asian patients exhibit distinct clinical phenotypes and risk profiles that may limit generalizability.^11^ Regional analyses reveal substantial ethnic differences in comorbidity patterns, with Malays and Indians demonstrating higher odds of coronary artery disease compared with Chinese populations, while Korean and Japanese patients show markedly lower rates.^11^ Despite representing a substantial proportion of the global heart failure burden, Asian populations remain underrepresented in studies examining health status determinants.^12^ Furthermore, older ambulatory patients are frequently excluded from clinical trials, creating uncertainty regarding factors influencing health status in this vulnerable demographic.^13^

This study aimed to identify and quantify independent predictors of health status among older ambulatory adults with chronic heart failure in an Asian population. We hypothesized that indicators of systemic burden, including functional class, comorbidity, frailty, and nutritional status, would predict KCCQ-12 scores independently of ejection fraction. We further examined whether these associations remained consistent across health status domains and heart failure phenotypes.

## Methods

### Study design and participants

This analytical observational, cross-sectional study followed Strengthening the Reporting of Observational Studies in Epidemiology (STROBE) guidelines for reporting.^14^ The source population comprised consecutive patients aged 60 years or older with chronic HF (diagnosed according to ESC guidelines for at least 3 months)^15^ attending Thong Nhat Hospital cardiology clinic between August 2024 and March 2025. Patients were excluded if they had acute decompensated heart failure (defined as hospitalization or urgent intravenous diuretic use within the previous 4 weeks) or severe cognitive impairment preventing informed consent or questionnaire completion. Sample size calculations used two approaches: first, estimating mean KCCQ-12 scores with 5-point precision^16^ (assuming a standard deviation of 25 and 95% confidence level) required 97 participants; second, Green’s formula for testing regression coefficients^17^ with six predictors (assuming a medium effect size, power of 0.80, and α = 0.05) required 110 participants. The final cohort of 150 participants exceeded both requirements. Ethical approval was obtained from the University of Medicine and Pharmacy at Ho Chi Minh City (2118/HDDD-DHYD) and Thong Nhat Hospital (121/2024/CN-BVTN-HDDD). The study complied with the Declaration of Helsinki,^18^ and all participants provided written informed consent before enrollment.

### Data collection and measurements

All procedures were completed during a single outpatient visit at Thong Nhat Hospital cardiology clinic. During the visit, trained staff conducted face-to-face interviews in Vietnamese and administered several validated instruments, including the Vietnamese KCCQ-12^16,19^ and the Clinical Frailty Scale (CFS),^20^ using structured case report forms developed for this study. Staff provided standardised instructions for all instruments and, where feasible, remained blinded to clinical data to minimise information bias. A cardiologist, blinded to the questionnaire results, performed clinical assessments and assigned NYHA functional class.^21^ Data for the Charlson Comorbidity Index (CCI), N-terminal pro–B-type natriuretic peptide (NT-proBNP) levels, and baseline LVEF were abstracted from medical records (selecting the most recent values within 3 months prior to the visit). Staff received protocol training with competency assessments. Data were entered into an electronic case report form with automated range and consistency checks. Analyses used anonymized data.

### Variable definitions

#### Primary and secondary outcomes

Participants completed a validated Vietnamese version of the KCCQ-12 during a baseline visit before clinical assessment. Scores for the four domains (physical limitation, symptom frequency, quality of life, and social limitation) were transformed to a 0–100 scale, with higher scores indicating better health. The primary outcome was the overall summary score, calculated as the mean of the available domain scores. The individual domain scores served as secondary outcomes. We prespecified a minimal clinically important difference of 5 or more points for the overall score, which is a conservative estimate consistent with published data.^22^

#### Key explanatory variables

LVEF was treated as a continuous variable in the primary regression model. Functional capacity was defined by the NYHA functional class, which we dichotomised for regression into mild (classes I–II) and advanced (classes III–IV) limitation.^21^ The CCI represented the comorbidity burden.^23^ While treated as a continuous variable in the primary regression model, for descriptive analyses, scores were stratified into low burden (scores 3–5), medium burden (6–7), and high burden (≥8). This categorisation was based on the distribution of scores within the study cohort rather than pre-specified a priori cut-points. Frailty was defined as a score of 5 or higher on the 9-point CFS. This threshold is widely used and validated for identifying at least mild frailty in older adults and is associated with adverse outcomes.

#### Other covariates

For descriptive purposes, we classified HF phenotype according to 2021 European Society of Cardiology guidelines based on LVEF: reduced (HFrEF; ≤40%), mildly reduced (HFmrEF; 41–49%), or preserved (HFpEF; ≥50%).^15^ In the primary model, LVEF was treated as a continuous variable. We recorded demographics (age, sex), lifestyle factors (smoking, alcohol use), and clinical measurements, including body mass index, resting heart rate and blood pressure (calculated as the mean of two measurements), and NT-proBNP concentrations. Polypharmacy was defined as using five or more concurrent medications.

We also counted the number of prescribed foundational HF drug classes (renin-angiotensin system inhibitors, beta-blockers, mineralocorticoid receptor antagonists, and sodium-glucose cotransporter-2 inhibitors; range 0–4). Functional status was assessed using the Katz Index of Independence in Activities of Daily Living,^24^ and nutritional status was evaluated with the Mini Nutritional Assessment Short-Form.^25^

#### Statistical analysis

Statistical analyses were performed using Stata version 19.5 (StataCorp, College Station, Texas, USA). Categorical variables were summarized as counts and percentages. Continuous variables were assessed for normality using the Shapiro-Wilk test and histogram inspection, then reported as mean with standard deviation or median with interquartile range.

Descriptive trends across ordered groups were tested using the Jonckheere-Terpstra test for continuous or ordinal outcomes and the Cochran-Armitage test for binary outcomes. KCCQ-12 summary and domain scores were compared across categories using the Mann-Whitney U test for two groups and the Jonckheere-Terpstra test for three or more ordered groups. The primary analysis used a multivariable linear regression model to identify predictors of the KCCQ-12 overall summary score. To separate chronological age from cumulative comorbidity, the model included age and the CCI as independent continuous predictors alongside LVEF, dichotomized NYHA functional class, binary frailty status, and sex. Results were expressed as adjusted regression coefficients with 95% confidence intervals using robust standard errors. Model diagnostics included residual plots for linearity and normality and variance inflation factors for multicollinearity.

Exploratory analyses stratified the multivariable regression model by HF phenotype (HFrEF, HFmrEF, and HFpEF) to examine predictor associations, with interaction p-values calculated to test for heterogeneity; however, these results are descriptive as the study was not powered for subgroup interactions. Secondary analyses evaluated the independent contribution of nutritional status (MNA-SF score) to health status and tested the robustness of findings through three sensitivity analyses: adjusting for log-transformed NT-proBNP to control for hemodynamic severity, excluding NYHA class to address potential circularity with patient-reported outcomes, and substituting frailty with nutritional status to assess systemic vulnerability across domains. All analyses utilized complete-case data; details regarding study exclusions due to missing data are provided in the enrolment flow diagram. A two-sided p-value less than 0.05 defined statistical significance, and no adjustments were made for multiple comparisons in exploratory analyses.

## Results

### Cohort and baseline description

The study cohort comprised 150 older adults with HF, categorized into three phenotype groups (Supplementary Figure 1). The median age was 75 years, with patients in the HFpEF group significantly older than those in the HFrEF group. Geriatric syndromes such as malnutrition, frailty, and limitations in daily activities were prevalent across the cohort. Most patients had moderate-to-high comorbidity burden and were on multiple medications. The proportion of patients with high comorbidity burden appeared to increase from the HFrEF to the HFpEF phenotype, though this trend was not statistically significant. Common comorbidities included hypertension, coronary artery disease, and dyslipidemia. Notably, the prevalence of atrial fibrillation increased significantly from the HFrEF to the HFpEF group (Table 1). Among patients with HFrEF, the prescription of foundational therapies was common. The majority were on combination therapy, with most receiving two to four different drug classes (Supplementary Table 1).

**Table 1.**
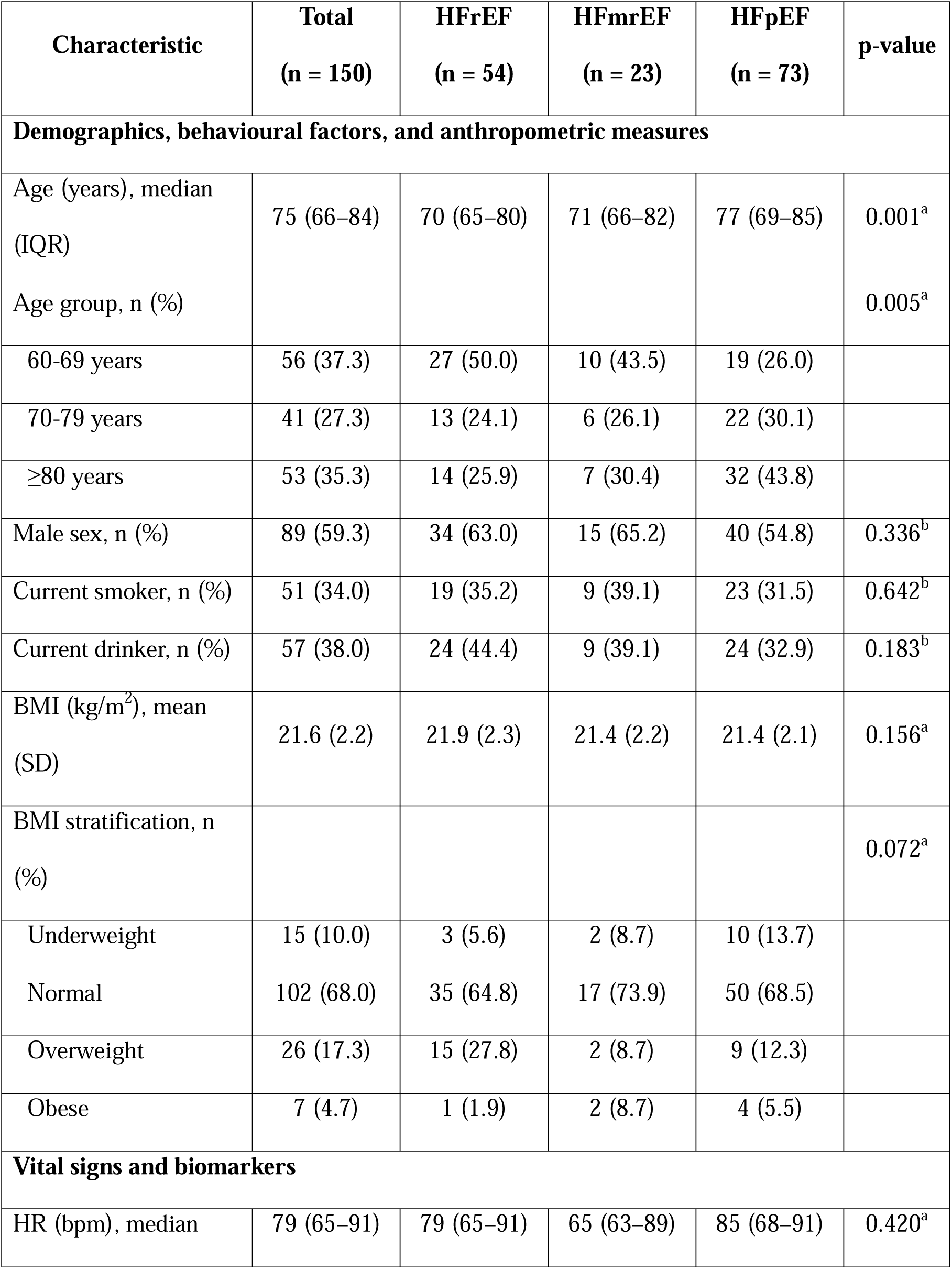
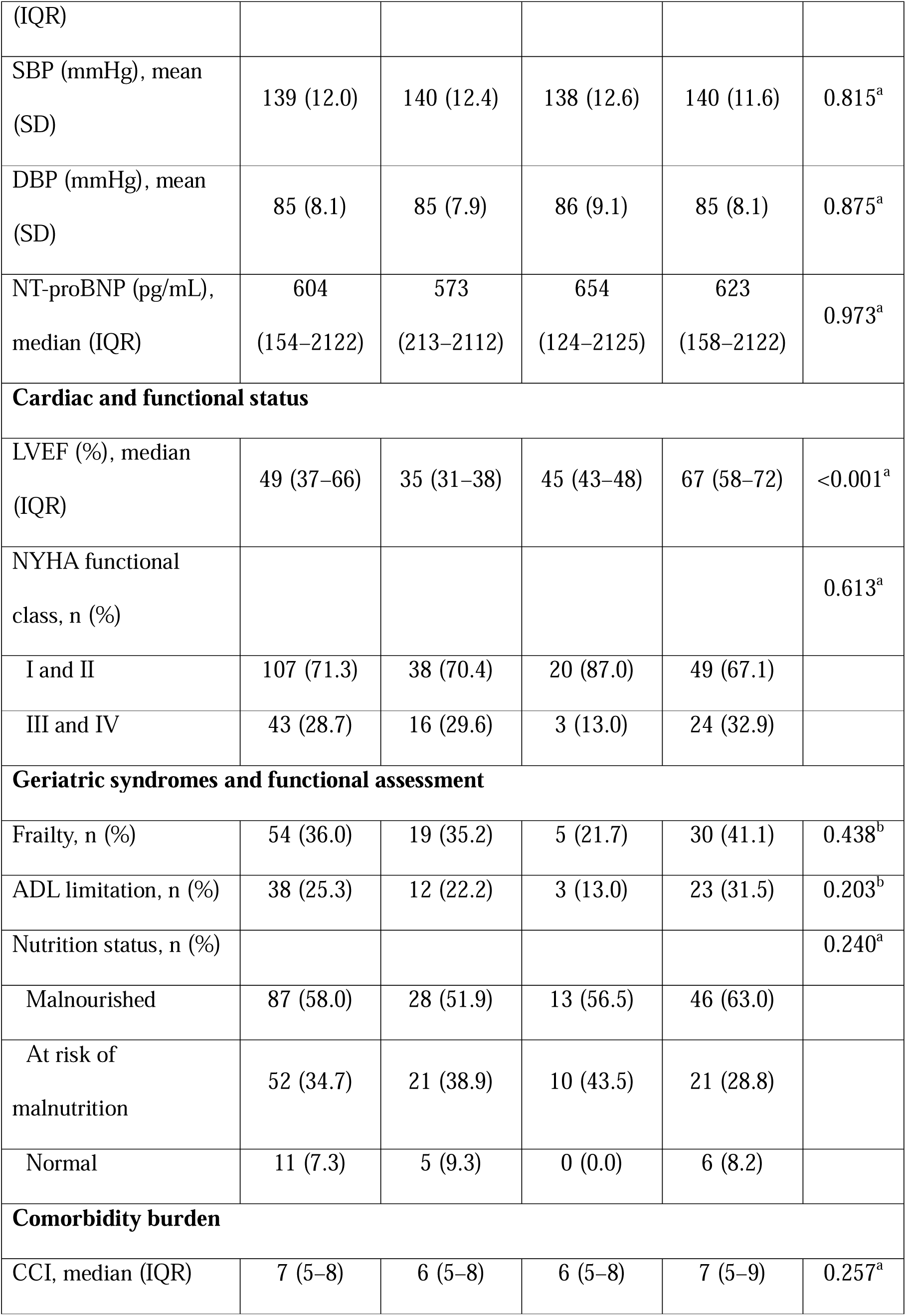
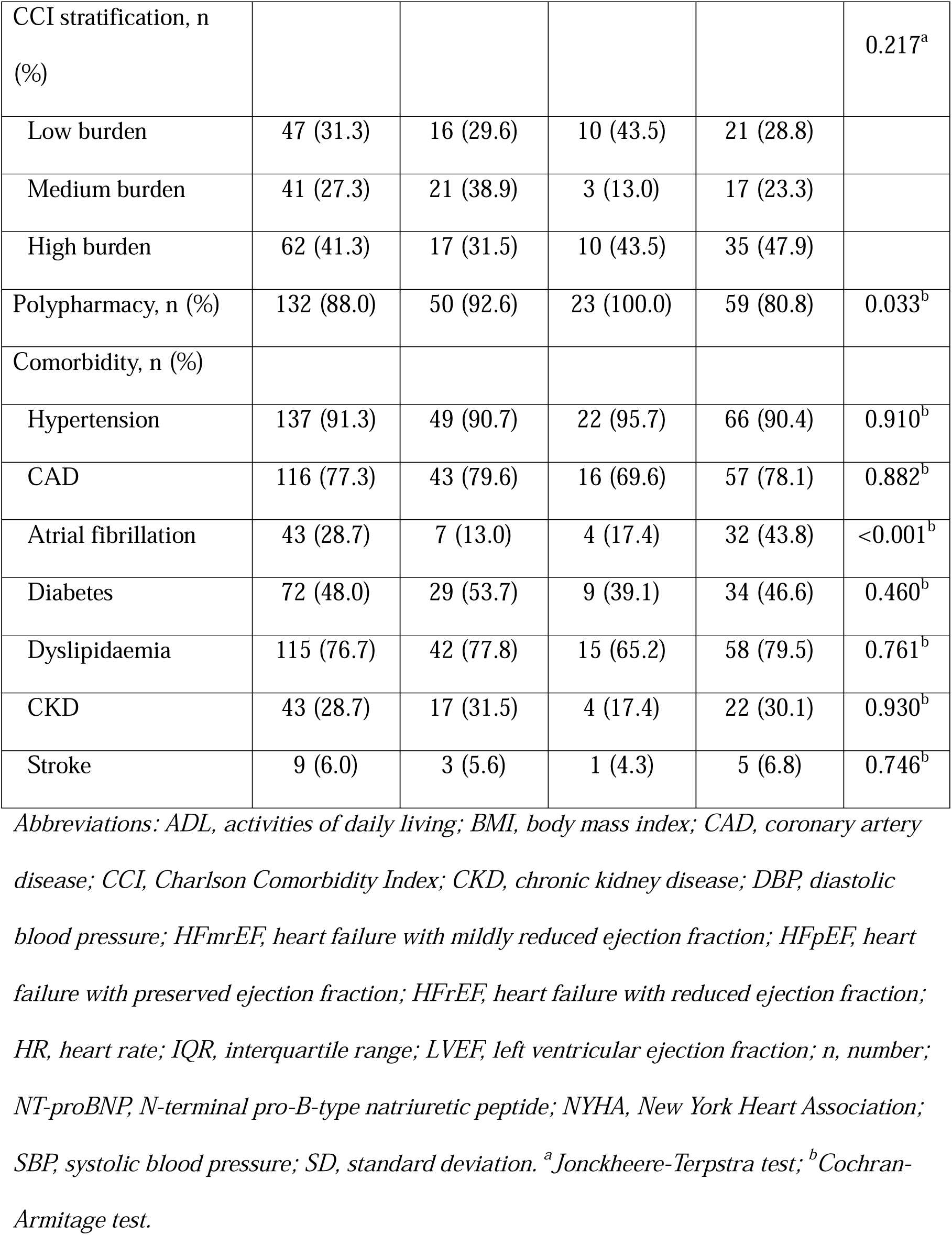

### Systemic burden and health status independent of ejection fraction

The median KCCQ-12 summary score was 68 and did not differ across HF phenotypes. Health status was poorer in patients with advanced NYHA functional class, frailty, higher comorbidity burden, and females (Table 2). A multivariable regression model explained 77% of variance in KCCQ-12 scores. Higher NYHA functional class, increased comorbidity burden, and frailty were significant predictors of poorer health status, while male sex was associated with higher scores (Table 3, Supplementary Figure 2). Age and LVEF were not significant predictors.

**Table 2.**
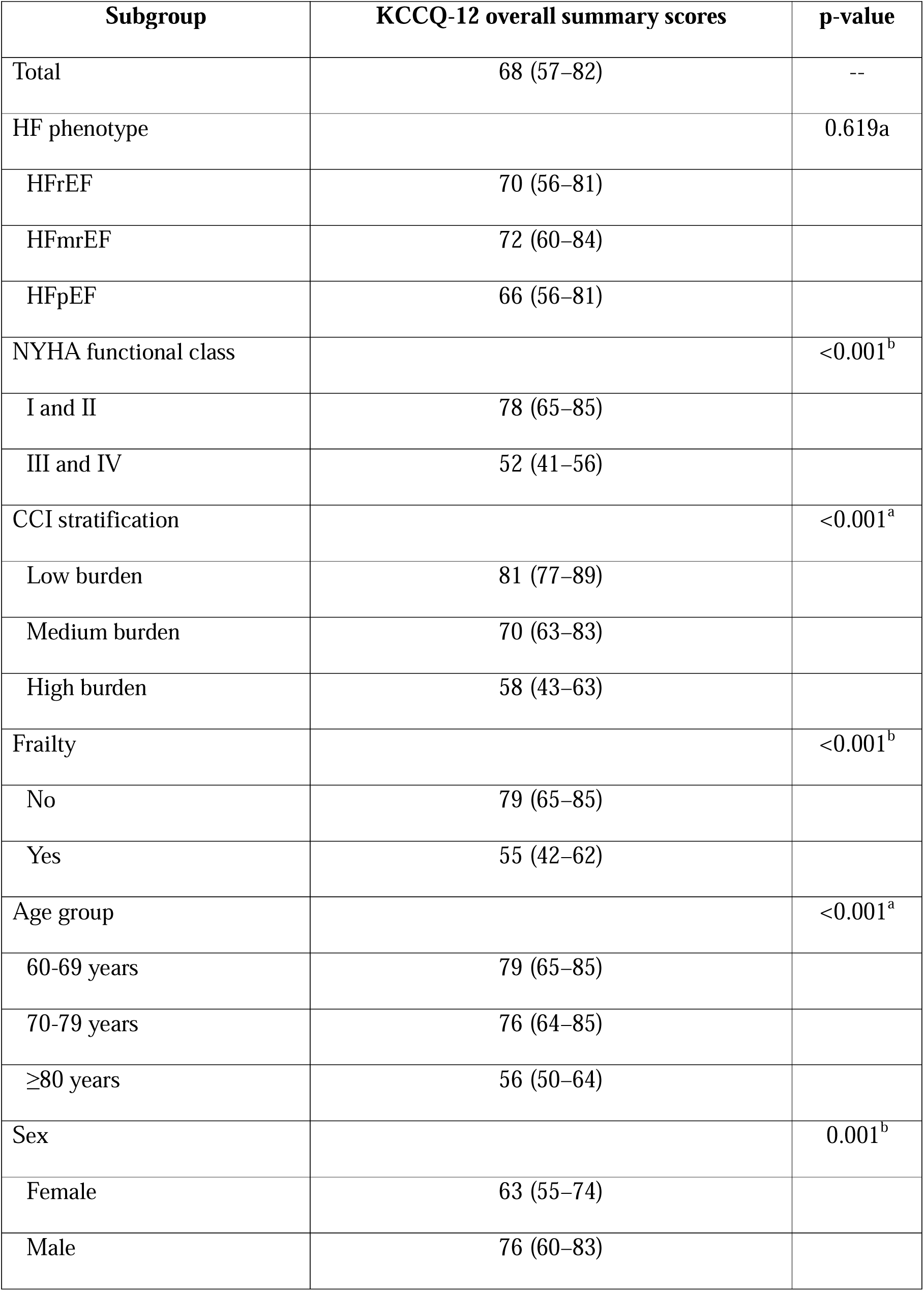
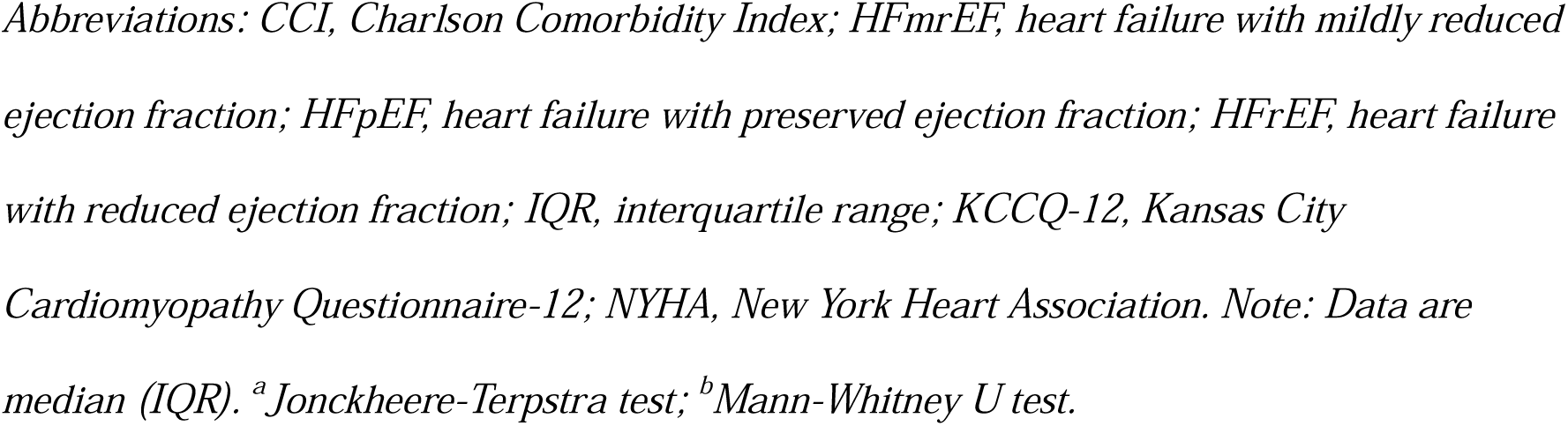

| **Subgroup** | **KCCQ-12 overall summary scores** | **p-value** |
| --- | --- | --- |
| Total | 68 (57–82) | -- |
| HF phenotype |  | 0.619<sup>a</sup> |
| HFrEF | 70 (56–81) |  |
| HFmrEF | 72 (60–84) |  |
| HFpEF | 66 (56–81) |  |
| NYHA functional class |  | <0.001<sup>b</sup> |
| I and II | 78 (65–85) |  |
| III and IV | 52 (41–56) |  |
| CCI stratification |  | <0.001<sup>a</sup> |
| Low burden | 81 (77–89) |  |
| Medium burden | 70 (63–83) |  |
| High burden | 58 (43–63) |  |
| Frailty |  | <0.001<sup>b</sup> |
| No | 79 (65–85) |  |
| Yes | 55 (42–62) |  |
| Age group |  | <0.001<sup>a</sup> |
| 60-69 years | 79 (65–85) |  |
| 70-79 years | 76 (64–85) |  |
| ≥80 years | 56 (50–64) |  |
| Sex |  | 0.001<sup>b</sup> |
| Female | 63 (55–74) |  |
| Male | 76 (60–83) |  |

*Abbreviations: CCI, Charlson Comorbidity Index; HFmrEF, heart failure with mildly reduced* *ejection fraction; HFpEF, heart failure with preserved ejection fraction; HFrEF, heart failure* *with reduced ejection fraction; IQR, interquartile range; KCCQ-12, Kansas City* *Cardiomyopathy Questionnaire-12; NYHA, New York Heart Association. Note: Data are* *median (IQR). <sup>a</sup>Jonckheere-Terpstra test; <sup>b</sup>Mann-Whitney U test.*

**Table 3.**

| **Total (n = 150)** |  |  |
| --- | --- | --- |
| **Predictor** | **Coefficient (95% CI)** | **p-value** |
| LVEF (per 1% increase) | 0.02 (-0.06 to 0.11) | 0.561 |
| NYHA functional class (III and IV vs I and II) | -15.88 (-19.03 to -12.73) | <0.001 |
| CCI (per 1 score increase) | -3.17 (-4.13 to -2.22) | <0.001 |
| Frailty (yes vs. no) | -6.82 (-10.31 to -3.32) | <0.001 |
| Age (per 1 increase) | -0.01 (-0.19 to 0.17) | 0.909 |
| Sex (male vs. female) | 3.21 (0.32 to 6.10) | 0.030 |
| Model statistics |  |  |
| R<sup>2</sup> | 0.767 |  |
| F-statistic | 67.94 | <0.001 |

*Abbreviations: CCI, Charlson Comorbidity Index; CI, confidence interval; LVEF, left* *ventricular ejection fraction; n, number; NYHA, New York Heart Association; ref, reference;* *VIF, variance inflation factor. Note: Diagnostic plots confirmed homoscedasticity, linearity,* *and residual normality; mean VIF (1.53) indicated no multicollinearity.*

### Systemic factors and health status domain scores

Consistent with the primary outcome, no significant trend was observed across HF phenotypes for any KCCQ-12 domain scores, including physical limitation, symptom frequency, quality of life, and social limitation. Subgroup analyses revealed consistent patterns of poorer health status, with patients in advanced NYHA functional classes, frail individuals, those with an increasing comorbidity burden, and females reporting significantly lower scores across all four domains (Table 4).

**Table 4.**

| **Predictor** | **Coefficient (95% CI)** | **p-value** |
| --- | --- | --- |
| LVEF (per 1% increase) | 0.02 (-0.07 to 0.10) | 0.717 |
| NYHA functional class (III and IV vs I and II) | -16.04 (-19.26 to -12.82) | <0.001 |
| CCI (per 1 score increase) | -3.09 (-4.03 to -2.14) | <0.001 |
| Frailty (yes vs. no) | -7.27 (-10.84 to -3.69) | <0.001 |
| Age (per 1 increase) | -0.01 (-0.21 to 0.18) | 0.878 |
| Sex (male vs. female) | 3.01 (0.15 to 5.87) | 0.039 |
| MNA-SF (per 1 point increase) | -0.35 (-0.76 to 0.06) | 0.096 |
| **Model statistics** |  |  |
| R<sup>2</sup> | 0.771 |  |
| F-statistic (7, 142) | 61.55 |  |

*Abbreviations: CCI, Charlson Comorbidity Index; CI, confidence interval; LVEF, left* *ventricular ejection fraction; MNA-SF, Mini Nutritional Assessment-Short Form; NYHA, New* *York Heart Association; VIF, variance inflation factor. Note: Diagnostic plots confirmed* *homoscedasticity, linearity, and residual normality; mean VIF (1.48) indicated no* *multicollinearity.*

### Nutritional status as independent predictor of health status

In the multivariable model evaluating nutritional status, the model explained 77% of variance in KCCQ-12 scores (Table 4). Nutritional status (MNA-SF) was not a significant independent predictor of health status after adjusting for frailty and other systemic factors. The established systemic predictors remained robust in this expanded model. Advanced NYHA functional class, increasing comorbidity burden, and the presence of frailty continued to significantly predict poorer health status. Male sex was associated with higher scores, while LVEF and age showed no significant association.

### Phenotype-specific associations between systemic factors and health status

Multivariable linear regression models stratified by HF phenotype explained a substantial proportion of variance in KCCQ-12 scores across all three groups (Table 5). In both HFrEF and HFpEF, more advanced NYHA class, frailty, and higher comorbidity burden were independent predictors of poorer health status. Among patients with HFpEF, male sex was also associated with higher scores. In the HFmrEF group, only higher comorbidity burden independently predicted lower health status. Health status declined more steeply with increasing comorbidities in HFmrEF than in HFrEF (Table 6, Table 7).

**Table 5.**

| Subgroup | Physical limitation<br>scores | p-value | Symptom<br>frequency<br>scores | p-value | Quality of life<br>scores | p-value | Social<br>limitation<br>scores | p-value |
| --- | --- | --- | --- | --- | --- | --- | --- | --- |
| Total | 67 (42–83) | -- | 73 (65–85) | -- | 50 (50–75) | -- | 75 (67–83) | -- |
| HF phenotype |  | 0.565 |  | 0.262 |  | 0.539 |  | 0.556 |
| HFrEF | 75 (42–75) |  | 74 (65–85) |  | 75 (50–75) |  | 71 (67–83) |  |
| HFmrEF | 67 (50–83) |  | 73 (70–94) |  | 50 (50–75) |  | 75 (67–92) |  |
| HFpEF | 58 (42–75) |  | 73 (60–79) |  | 50 (50–75) |  | 67 (67–83) |  |
| NYHA functional class |  | <0.001 |  | <0.001 |  | <0.001 |  | <0.001 |
| I and II | 75 (58–83) |  | 79 (70–92) |  | 75 (50–75) |  | 75 (67–92) |  |
| III and IV | 42 (25–42) |  | 54 (42–67) |  | 50 (38–50) |  | 58 (50–67) |  |
| CCI stratification |  | <0.001 |  | <0.001 |  | <0.001 |  | <0.001 |
| Low burden | 83 (75–100) |  | 85 (73–94) |  | 75 (50–88) |  | 83 (75–92) |  |
| Medium burden | 67 (58–75) |  | 73 (67–85) |  | 75 (50–75) |  | 75 (67–83) |  |
| High burden | 42 (33–58) |  | 65 (50–73) |  | 50 (50–50) |  | 67 (50–67) |  |
| Frailty |  | <0.001 |  | <0.001 |  | <0.001 |  | <0.001 |
| No | 75 (58–83) |  | 79 (70–92) |  | 75 (50–88) |  | 75 (67–92) |  |
| Yes | 42 (33–58) |  | 60 (44–70) |  | 50 (50–50) |  | 63 (50–67) |  |
| Age group |  | <0.001 |  | <0.001 |  | <0.001 |  | <0.001 |
| 60-69 years | 75 (58–83) |  | 79 (70–93) |  | 75 (50–75) |  | 83 (67–92) |  |
| 70-79 years | 75 (58–83) |  | 73 (67–92) |  | 75 (50–88) |  | 75 (67–83) |  |
| ≥80 years | 42 (42–58) |  | 65 (50–70) |  | 50 (50–75) |  | 67 (58–67) |  |
| Sex |  | 0.011 |  | 0.001 |  | 0.002 |  | <0.001 |
| Female | 58 (42–75) |  | 70 (58–73) |  | 50 (50–75) |  | 67 (58–75) |  |
| Male | 75 (50–83) |  | 75 (67–92) |  | 75 (50–75) |  | 75 (67–83) |  |

Abbreviations: CCI, Charlson Comorbidity Index; HFmrEF, heart failure with mildly reduced ejection fraction; HFpEF, heart failure with preserved ejection fraction; HFrEF, heart failure with reduced ejection fraction; IQR, interquartile range; KCCQ-12, Kansas City Cardiomyopathy Questionnaire-12; NYHA, New York Heart Association. Note: Data are median (IQR).

**Table 6.**

|  | **HFrEF (n = 54)** |  | **HFmrEF (n = 23)** |  | **HFpEF (n = 73)** |  |
| --- | --- | --- | --- | --- | --- | --- |
| **Predictor** | **Coefficient (95% CI)** | **p-value** | **Coefficient (95% CI)** | **p-value** | **Coefficient (95% CI)** | **p-value** |
| NYHA functional class (III and IV vs. I and II) | -18.18 (-23.94 to -12.41) | <0.001 | -14.95 (-38.43 to 8.54) | 0.197 | -17.30 (-21.24 to -13.34) | <0.001 |
| CCI (per 1 score increase) | -2.25 (-3.66 to -0.84) | 0.002 | -7.00 (-10.35 to -3.65) | <0.001 | -2.80 (-4.27 to -1.32) | <0.001 |
| Frailty (yes vs. no) | -9.61 (-15.29 to -3.92) | 0.001 | -0.80 (-14.60 to 13.00) | 0.904 | -6.07 (-11.01 to -1.13) | 0.017 |
| Age (per 1 increase) | 0.04 (-0.23 to 0.31) | 0.768 | 0.31 (-0.46 to 1.09) | 0.405 | -0.02 (-0.31 to 0.27) | 0.897 |
| Sex (male vs. female) | 0.45 (-4.77 to 5.66) | 0.864 | 0.36 (-8.41 to 9.12) | 0.933 | 6.21 (1.89 to 10.53) | 0.006 |
| **Model statistics** |  |  |  |  |  |  |
| R<sup>2</sup> | 0.800 |  | 0.713 |  | 0.791 |  |
| F-statistic | 32.62 |  | 10.74 |  | 43.81 |  |

Abbreviations: CCI, Charlson Comorbidity Index; CI, confidence interval; HFmrEF, heart failure with mildly reduced ejection fraction; HFpEF,
heart failure with preserved ejection fraction; HFrEF, heart failure with reduced ejection fraction; n, number; NYHA, New York Heart
Association; VIF, variance inflation factor. Note: Diagnostic plots confirmed homoscedasticity, linearity, and residual normality for all
subgroups; mean VIFs (HFrEF 1.61; HFmrEF 2.68; HFpEF 1.78) indicated no multicollinearity.

**Table 7.**

| **Predictor** | **Interaction p-value** |
| --- | --- |
| NYHA functional class | 0.766 |
| CCI | 0.005 |
| Frailty | 0.198 |
| Age | 0.464 |
| Sex | 0.985 |

*Abbreviations: CCI, Charlson Comorbidity Index; NYHA, New York Heart Association.*

### Systemic predictors independent of NT-proBNP levels

In the sensitivity analysis incorporating log-transformed NT-proBNP to adjust for hemodynamic stress, model fit improved minimally compared to the primary model (Supplementary Table 2). NT-proBNP levels were not significantly associated with KCCQ-12 scores. The associations between systemic factors and health status remained consistent.

Higher NYHA functional class, greater comorbidity burden, and frailty remained significant independent predictors of lower health status scores. LVEF was not significantly associated with health status. Model assumptions of homoscedasticity, linearity, and residual normality were satisfied.

### Objective systemic factors excluding functional class

In the sensitivity analysis excluding NYHA functional class, the model explained 66% of variance in KCCQ-12 scores, reduced from the primary model (Supplementary Table 3). Systemic factors remained significant predictors. The association between frailty and health status was larger in this model compared to the primary analysis. Greater comorbidity burden remained significantly associated with lower health status scores. Male sex was associated with higher scores, while LVEF and age were not significantly associated with health status.

### Nutritional status substitution for frailty assessment

In the sensitivity analysis replacing frailty with nutritional status, the model explained 75% of variance in KCCQ-12 scores. Nutritional status (MNA-SF) was not significantly associated with health status (Supplementary Table 4). Other systemic predictors remained significant, including higher NYHA functional class and greater comorbidity burden, both associated with lower health status scores. LVEF and age were not significantly associated with health status. Male sex was also not significantly associated with health status in this model.

## Discussion

### Summary of key findings

This study examined health status determinants in 150 consecutively enrolled older Vietnamese adults with chronic heart failure (median age 75 years) using the Kansas City Cardiomyopathy Questionnaire-12. Multivariable regression analysis demonstrated that geriatric syndromes, rather than cardiac-specific indices, determined patient-reported health status. Frailty (Clinical Frailty Scale score of 5 or higher), comorbidity burden (Charlson Comorbidity Index), and functional capacity (New York Heart Association class) independently predicted KCCQ-12 scores, collectively explaining approximately 77% of variance. Left ventricular ejection fraction and N-terminal pro-B-type natriuretic peptide showed no significant independent association with health status across all heart failure phenotypes. Nutritional status correlated with health status in univariate analyses but was subsumed by frailty in multivariable models, indicating frailty integrates catabolic decline, functional loss, and nutritional vulnerability. Male patients reported consistently better health status scores than females independent of clinical severity. These findings support a geriatric cardiology model prioritizing functional preservation and holistic assessment over isolated cardiac metrics for older Asian adults.

### The discordance between hemodynamics and health status

The absence of statistical association between left ventricular ejection fraction or N-terminal pro-B-type natriuretic peptide and KCCQ-12 scores challenges the conventional assumption that symptom severity reflects cardiac dysfunction. Symptom burden in older adults is primarily driven by peripheral factors including skeletal muscle dysfunction, endothelial abnormalities, and impaired oxygen extraction rather than central hemodynamic inadequacy.^26^ Chronic heart failure induces skeletal myopathy characterized by fiber-type shifts, mitochondrial dysfunction, and reduced capillary density.^27^ In Asian populations with lower baseline muscle mass, progression to sarcopenia occurs earlier and more severely, leading to early anaerobic metabolism and muscle ergoreflex activation during minimal exertion.^28^ Consequently, patients may demonstrate improved left ventricular ejection fraction via guideline-directed medical therapy yet remain symptomatic if peripheral metabolic capacity remains compromised. Additionally, left ventricular ejection fraction is a load-dependent volumetric measure that fails to capture longitudinal strain, diastolic relaxation abnormalities, or left atrial mechanics.^29^

N-terminal pro-B-type natriuretic peptide, while indicating wall tension and volume overload, does not reflect functional adaptation or psychosocial resilience. Older adults with chronically elevated filling pressures often adapt by restricting physical activity to avoid dyspnea, reporting fewer symptoms despite elevated biomarker levels. Furthermore, N-terminal pro-B-type natriuretic peptide levels are influenced by renal dysfunction, atrial fibrillation, anemia, and age independent of heart failure severity.^30^ The decoupling of central hemodynamics from symptoms supports the peripheral hypothesis, suggesting that interventions targeting skeletal muscle and metabolic competence, such as resistance training and nutritional supplementation, may improve health status more effectively than further titration of hemodynamically active medications in older Asian adults.

### The dominant role of frailty and geriatric syndromes

Frailty is a central, independent determinant of health status in older adults with heart failure. The strong association between Clinical Frailty Scale scores and KCCQ-12 domains illustrates that frailty amplifies symptom burden beyond the effects of age or comorbidity alone. Frailty represents a multidimensional physiological vulnerability involving inflammation, sarcopenia, and loss of homeostatic reserve that is distinct from chronological aging or the presence of multiple diseases.^31^ The prevalence of frailty in this cohort was 36 percent, emphasizing the significance of this syndrome even in populations with lower BMI, where sarcopenia and functional decline can be masked by a lean phenotype.

While nutritional status showed significance in univariate analyses, its predictive value was subsumed by frailty in multivariable models. Frailty integrates nutritional decline with cognitive impairment, physical limitations, and comorbidities, making it a superior tool for rapid risk stratification in outpatient settings.^32^ However, nutritional screening remains valuable to identify reversible contributors to frailty. The high impact of frailty in Vietnamese patients may reflect regional factors including limited access to preventive geriatric care and lifelong nutritional and infectious exposures, which accelerate onset and severity. In Asian settings, familial dependency magnifies the functional and social consequences of frailty.

Integrating frailty into routine assessment, and adjusting management intensity based on frailty severity, is critical. Robust patients benefit from guideline-directed therapy, while frail patients require geriatric-focused interventions, including physical rehabilitation, deprescribing, and individualized care planning.^15^

### Demographic and phenotypic nuances

Male sex was associated with health status scores approximately 3 points higher than females after adjusting for other factors. Biological explanations include greater skeletal muscle mass providing metabolic reserve, higher hemoglobin levels, and reduced diastolic stiffness in men.^33^ However, sociocultural factors likely contribute substantially in Asian populations, where older men may underreport symptoms due to cultural stoicism while women experience greater physical and emotional strain from dual roles as patients and caregivers.

Women demonstrate higher rates of depression and anxiety in heart failure, which independently suppresses health status scores.^34^ The female disadvantage in heart failure with preserved ejection fraction is particularly pronounced, as older women experience disproportionate symptomatic intolerance due to arterial stiffening and ventricular-arterial uncoupling.^34^

Heart failure with mildly reduced ejection fraction displayed a distinct profile where comorbidity burden was the dominant health status predictor, with a regression coefficient more than double that of reduced or preserved ejection fraction phenotypes. This suggests symptoms in mildly reduced ejection fraction result from cumulative mild organ dysfunctions rather than isolated cardiac impairment. Effective quality of life improvement may require aggressive non-cardiac comorbidity management including diabetes control, anemia correction, and pulmonary disease treatment rather than focusing solely on heart failure pharmacotherapy.

The high multimorbidity burden (median Charlson Comorbidity Index of 7) reflects Southeast Asia’s epidemiological transition, where heart failure populations exhibit complex admixtures of hypertension, diabetes, infectious disease histories, and lifestyle diseases.^35^ The exceptionally high burden of diabetes and chronic kidney disease necessitates multimorbidity-focused care plans rather than disease-specific guidelines.

### Strengths and limitations

This study addresses gaps in heart failure literature by enrolling consecutive older adults (aged 60 years or older) in a real-world Vietnamese ambulatory setting, a population underrepresented in North American and European trials. Simultaneous collection of clinician-reported, patient-reported, and objective measures enables comprehensive assessment of health status determinants. The Vietnamese-validated KCCQ-12 ensures cultural and linguistic appropriateness. Explicit inclusion of frailty and nutritional assessment bridges cardiology and geriatrics. Robust associations despite modest sample size reflect substantial effect sizes.

Sample size limited subgroup analysis power, particularly for mildly reduced ejection fraction. Cross-sectional design precludes causal inference regarding frailty and health status. Survivorship and referral bias may exist, as ambulatory patients may be healthier than homebound individuals, potentially underestimating frailty impact. Unmeasured variables including socioeconomic status, health literacy, family support, social isolation, and depression may contribute to residual confounding. Nevertheless, robustness of core associations across sensitivity analyses supports reliability and generalizability to similar urban Asian populations.

### Future perspectives and clinical implications

These findings support routine frailty screening using the Clinical Frailty Scale in Asian heart failure clinics. Patient-reported outcomes should replace left ventricular ejection fraction as therapeutic success metrics in older adults. International guidelines should mandate frailty assessment and incorporate patient-reported outcomes as primary endpoints for Asian populations.^3^ Future research must test frailty-targeted interventions, including nutritional supplementation and resistance training, through randomized trials. Longitudinal studies examining social determinants and multigenerational cohabitation impacts on frailty progression are essential for developing effective community-based interventions.

## Conclusion

This study demonstrates that left ventricular ejection fraction and N-terminal pro-B-type natriuretic peptide do not predict health status in older Asian adults with heart failure.

Geriatric syndromes including frailty, multimorbidity, and functional limitation determine patient-reported outcomes. These findings support integrating geriatric principles into cardiovascular practice through systematic frailty assessment and functional preservation. Optimizing heart failure care in aging Asian populations requires holistic, multidisciplinary approaches addressing frailty with equivalent rigor as hemodynamic management.

## Author contributions

All authors approved the final submitted version and any revised versions. Nguyen Le Huy Hoang is the guarantor with full data access and responsibility for accuracy.

## Data Availability

All data produced in the present study are available upon reasonable request to the corresponding author. Due to ethical and privacy considerations related to patient confidentiality, individual-level data cannot be publicly deposited but may be shared following institutional review and appropriate data use agreements.

## Acknowledgements

We thank the patients and their families for participating in this study. We are grateful to the clinicians, echocardiography technologists, and nursing staff at Thong Nhat Hospital for their support with patient recruitment, clinical assessments, and echocardiographic evaluations. We acknowledge the use of the KCCQ-12, licensed from CV Outcomes, Inc. (Kansas City, Missouri, USA).

