## Supplementary Material for "Independent contributions of functional class, comorbidity, and frailty to health status in elderly adults with heart failure"

Supplementary Figure 1

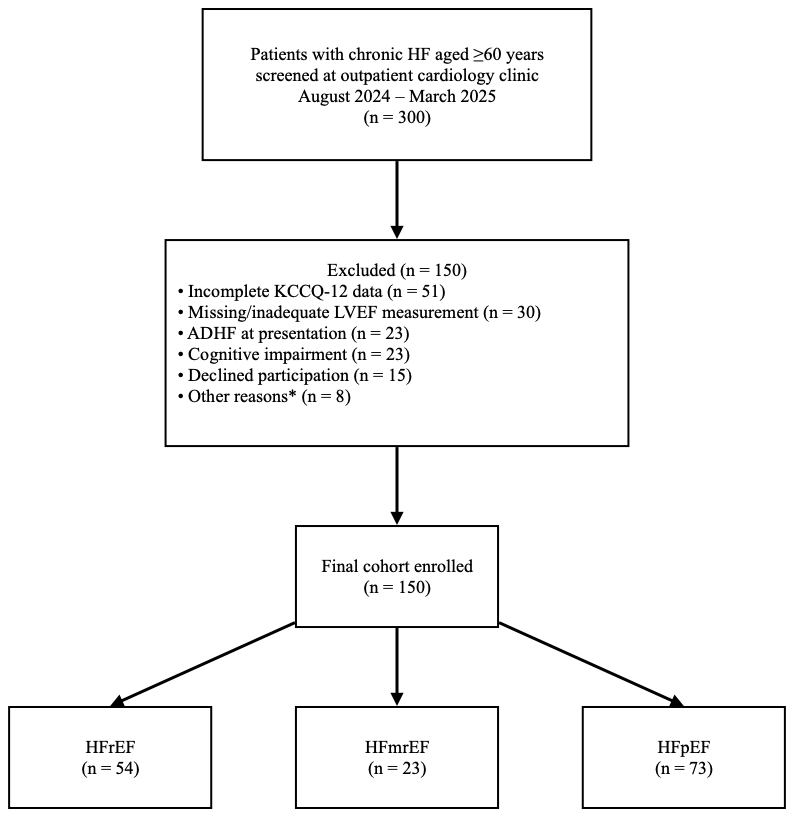
STROBE-style diagram illustrating the screening and enrolment process. Of 300 patients screened, 150 were excluded for reasons including incomplete questionnaire data (n = 51), missing left ventricular ejection fraction measurement (n = 30), acute decompensated heart failure (n = 23), cognitive impairment (n = 23), and declined participation (n = 15), resulting in a final cohort of 150 patients

*Other reasons for exclusion included severe non-cardiac illness (n = 5) and planned relocation (n = 3).

Abbreviations: ADHF, acute decompensated heart failure; HFmrEF, heart failure with mildly reduced ejection fraction; HFpEF, heart failure with preserved ejection fraction; HFrEF, heart failure with reduced ejection fraction.

Supplementary Table 1

| Variable | HFrEF  **(n = 54)** |
| --- | --- |
| Drug class | |
| RAASi | 37 (68.5%) |
| BB | 28 (51.9%) |
| MRA | 48 (88.9%) |
| SGLT2i | 42 (77.8%) |
| Number of foundational drugs | |
| 0 | 2 (3.7%) |
| 1 | 1 (1.9%) |
| 2 | 15 (27.8%) |
| 3 | 20 (37.0%) |
| 4 | 16 (29.6%) |

*Abbreviations: BB, beta-blocker; HFrEF, heart failure with reduced ejection fraction; MRA, mineralocorticoid receptor antagonist; n, number; RAASi, renin-angiotensin-aldosterone system inhibitor; SGLT2i, sodium-glucose cotransporter-2 inhibitor.*

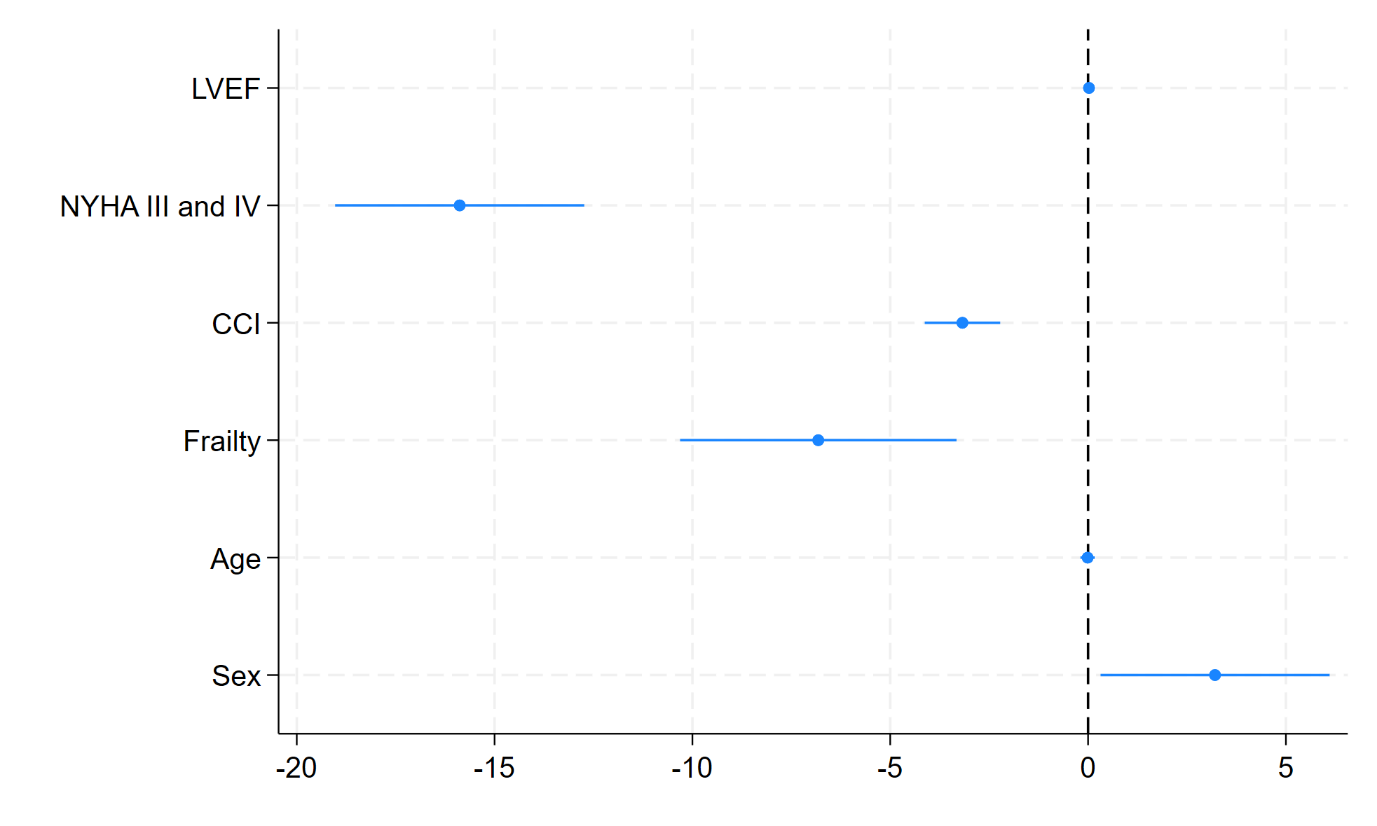
Supplementary Figure 2

The plot shows the adjusted regression coefficients (blue dots) and 95% confidence intervals (horizontal lines) from the primary multivariable linear regression model (n = 150). The dashed vertical line indicates a coefficient of zero, representing no effect. Coefficients and confidence intervals that do not cross this line are statistically significant.

Abbreviations: CCI, Charlson Comorbidity Index; LVEF, left ventricular ejection fraction; NYHA, New York Heart Association.

Supplementary Table 2

| **Predictor** | **Coefficient (95% CI)** | **p-value** |
| --- | --- | --- |
| LVEF (per 1% increase) | 0.02 (-0.06 to 0.11) | 0.586 |
| NYHA functional class (III and IV vs I and II) | -16.11 (-19.30 to -12.92) | <0.001 |
| CCI (per 1 score increase) | -3.17 (-4.11 to -2.22) | <0.001 |
| Frailty (yes vs. no) | -6.74 (-10.20 to -3.28) | <0.001 |
| Age (per 1 increase) | -0.01 (-0.19 to 0.18) | 0.944 |
| Sex (male vs. female) | 3.26 (0.36 to 6.16) | 0.028 |
| log(NT-proBNP) (per 1 increase) | -0.33 (-1.14 to 0.48) | 0.419 |
| **Model statistics** |  |  |
| R^2^ | 0.768 |  |
| F-statistic (7, 142) | 58.84 |  |

*Abbreviations: CCI, Charlson Comorbidity Index; CI, confidence interval; LVEF, left ventricular ejection fraction; NT-proBNP, N-terminal pro-B-type natriuretic peptide; NYHA, New York Heart Association; VIF, variance inflation factor. Note: Diagnostic plots confirmed homoscedasticity, linearity, and residual normality; mean VIF (1.47) indicated no multicollinearity.*

Supplementary Table 3

| **Predictor** | **Coefficient (95% CI)** | **p-value** |
| --- | --- | --- |
| LVEF (per 1% increase) | 0.04 (-0.07 to 0.14) | 0.511 |
| CCI (per 1 score increase) | -3.82 (-4.96 to -2.67) | <0.001 |
| Frailty (yes vs. no) | -11.56 (-16.10 to -7.03) | <0.001 |
| Age (per 1 increase) | -0.17 (-0.42 to 0.08) | 0.173 |
| Sex (male vs. female) | 4.12 (0.52 to 7.72) | 0.025 |
| **Model statistics** |  |  |
| R^2^ | 0.659 |  |
| F-statistic (5, 144) | 54.38 |  |

*Abbreviations: CCI, Charlson Comorbidity Index; CI, confidence interval; LVEF, left ventricular ejection fraction; VIF, variance inflation factor. Note: Diagnostic plots confirmed homoscedasticity, linearity, and residual normality; mean VIF (1.46) indicated no multicollinearity.*

Supplementary Table 4

| **Predictor** | **Coefficient (95% CI)** | **p-value** |
| --- | --- | --- |
| LVEF (per 1% increase) | 0.03 (-0.06 to 0.12) | 0.508 |
| NYHA functional class (III and IV vs I and II) | -18.01 (-21.49 to -14.54) | <0.001 |
| CCI (per 1 score increase) | -3.93 (-4.73 to -3.13) | <0.001 |
| Age (per 1 increase) | -0.02 (-0.22 to 0.19) | 0.856 |
| Sex (male vs. female) | 3.02 (-0.01 to 6.06) | 0.051 |
| MNA-SF (per 1 point increase) | -0.25 (-0.65 to 0.15) | 0.223 |
| **Model statistics** |  |  |
| R^2^ | 0.750 |  |
| F-statistic (6, 143) | 66.47 |  |

*Abbreviations: CCI, Charlson Comorbidity Index; CI, confidence interval; LVEF, left ventricular ejection fraction; MNA-SF, Mini Nutritional Assessment-Short Form; NYHA, New York Heart Association; VIF, variance inflation factor. Note: Diagnostic plots confirmed homoscedasticity, linearity, and residual normality; mean VIF (1.28) indicated no multicollinearity.*
